# Factors shaping vaginal microbiota long-term community dynamics in young adult women

**DOI:** 10.1101/2024.04.08.24305448

**Authors:** Tsukushi Kamiya, Nicolas Tessandier, Baptiste Elie, Claire Bernat, Vanina Boué, Sophie Grasset, Soraya Groc, Massilva Rahmoun, Christian Selinger, Michael S. Humphrys, Marine Bonneau, Christelle Graf, Vincent Foulongne, Jacques Reynes, Vincent Tribout, Michel Segondy, Nathalie Boulle, Jacques Ravel, Carmen Lía Murall, Samuel Alizon

## Abstract

The vaginal microbiota is known to affect women’s health. Yet, there is a notable paucity of high-resolution follow-up studies lasting several months, which would be required to interrogate the long-term dynamics and associations with demographic and behavioural covariates. Here, we present a high-resolution longitudinal cohort of 125 women followed for a median duration of 8.6 months, providing 11 samples per woman. Using a hierarchical Bayesian Markov model, we characterised the patterns of vaginal microbiota community persistence and transition, simultaneously estimated the impact of 16 covariates and quantified individual variability among women. We showed that ‘optimal’ (Community State Type (CST) I, II, and V) and ‘sub-optimal’ (CST III) communities are more stable over time than ‘non-optimal’ (CST IV) ones. Furthermore, we found that some covariates — most notably alcohol consumption — impacted the probability of shifting from one CST to another. We performed counterfactual simulations to confirm that alterations of key covariates, such as alcohol consumption, could shape the prevalence of different microbiota communities in the population. Finally, our analyses indicated that there is a relatively canalised pathway leading to the deterioration of vaginal microbiota communities, whereas the paths to recovery can be highly individualised among women. In addition to providing one of the first insights into vaginal microbiota dynamics over a year, our study showcases a novel application of a hierarchical Bayesian Markov model to clinical cohort data with many covariates. Our findings pave the way for an improved mechanistic understanding of microbial dynamics in the vaginal environment and the development of novel preventative and therapeutic strategies to improve vaginal health.

## Introduction

Epithelia of the human body are host to a diverse array of microorganisms. These microorganisms are collectively referred to as microbiota and their compositions are tightly associated with human health. In the human vaginal environment, the description of the microbiota dates back to Albert Döderlein in 1892. Its composition has been demonstrated to impact the acquisition risk of several sexually transmitted infections (STIs) [1], fertility (especially in medically-assisted procreation procedures) [2], and general well-being [3].

Vaginal microbiota communities comprise hundreds of species. To facilitate understanding, the variation in community composition is usually reduced to a handful of categories that capture key compositional signatures, such as the dominance of certain species or species evenness. This dimensionality reduction filters out noise in the data and facilitates the identification and visualisation of key patterns and relationships.

Potential drawbacks of reducing continuous variation include the risk of losing subtle but meaningful signals within the microbiota, as less dominant or rare taxa may be excluded despite their potential importance. Compared to the gut microbiota, however, vaginal microbiota communities tend to be highly structured and are often dominated by a small handful of species whose functional ecology is well-documented [4]. This contrasts with the highly diverse gut microbiota, where defining discrete community types, such as “enterotypes,” remains contentious [5]. The high diversity and evenness in gut microbiota introduce continuous variations that can be oversimplified by strict categorical clustering. In contrast, vaginal microbiota composition aligns more naturally with categorical clustering, providing a robust understanding of key microbial patterns without significantly sacrificing interpretability.

One dimensionality reduction framework, i.e., community state types (CSTs), introduced by Ravel et al. [6], categorises vaginal microbial communities into five discrete state types. The CSTs considered ‘optimal’ for health are dominated by *Lactobacillus* species; *Lactobacillus crispatus*, *L. gasseri*, and *L. jensenii* for CST I, II, and V, respectively. Lactobacilli produce lactic acid and hydrogen peroxide, which create an acidic environment that helps to inhibit the growth of harmful pathogens [7]. On the other end of the spectrum, CST IV is the primary microbial context of bacterial vaginosis (BV), which elevates the risk of STI acquisition and spontaneous preterm birth, and is associated with symptoms such as malodor, discharge, and itching [4, 8]. This community is characterised by a diverse assemblage of anaerobic bacterial species from the *Gardnerella*, *Prevotella*, and *Fannyhessea* genera: recent classifications include sub-categories within CST IV (i.e., IV-A, IV-B, IV-C), each with a distinct microbial profile [9]. Finally, CST III, characterised by a dominance of *L. iners*, is considered ‘sub-optimal’ for women’s health. While *L. iners* is a member of the *Lactobacillus* genus, it is less effective at producing lactic acid and hydrogen peroxide. As such, women with CST III tend to exhibit higher vaginal pH than those with CST I and are more prone to experiencing adverse health consequences, including vaginal infections [10].

The CST classification represents a snapshot of the microbiota community at the time of sampling that facilitates the examination of clinically relevant microbiota variations across time and women. The development of the modern pipeline — through meta-barcoding sequencing of 16S DNA and clustering algorithms [9] — allows for CST-typing with enhanced efficiency and reduced observer bias compared to conventional microscopy-based methods of vaginal microbiota community typing (e.g., Nugent score).

The composition of vaginal microbiota is characteristically variable over both short and long timescales [11]. For instance, vaginal microbiota shifts throughout a woman’s life, with prepubescent girls and postmenopausal women exhibiting lower levels of *Lactobacillus* dominance compared to women of reproductive age, though their bacterial communities are distinct from the CST IV typically seen during reproductive years [4]. On a short timescale, daily CST fluctuations are observed in some women of reproductive age, while others remain remarkably stable across menstrual cycles, suggesting that diverse factors influence the dynamics of vaginal microbiota communities [12]. For example, menstruation is a key driver of monthly dynamics, while clinical interventions such as antibiotics and probiotics can cause temporary perturbations [4].

A notable gap in the existing literature remains in the understanding of the long-term dynamics of vaginal microbiota in reproductive-aged women across several months. While some studies do follow this timespan, they focus on pregnancy-specific dynamics [13, 14], have large intervals between samples (often exceeding three months) [15], or involve modest sample sizes [16]. These limitations hinder our ability to fully understand the long-term patterns of CST stability and transitions in the general population of reproductive-aged women, and the influence of clinically relevant factors such as demography, lifestyle, sexual practices, and medication.

In this study, we introduce an original follow-up cohort of 125 women in Montpellier, France. Our cohort presents a high-resolution longitudinal follow-up study with 2,103 microbial samples, spanning a median duration of over 8.6 months and a median of 11 samples per woman. We devise a hierarchical Bayesian Markov model to estimate transition probabilities between CSTs, associations between the transitions and 16 relevant covariates, and individual variability among women.

## Materials and Methods

### Longitudinal clinical data

The samples originated from the PAPCLEAR monocentric longitudinal cohort study, which followed 189 women longitudinally between 2016 and 2020. The participants were recruited through posters and leaflets circulated at the main sexually transmitted infection detection centre (CeGIDD) at the University Hospital of Montpellier (CHU) and at and around university campuses in the city. The inclusion criteria were to be between 18 and 25 years old, to be living in the area of Montpellier, France, to be in good health (no chronic disease), not to have a history of human papillomavirus (HPV) infection (e.g., genital warts or highgrade cervical lesion), and to report at least one new sexual partner over the last 12 months. Additional details about the protocol can be found elsewhere [17]. The longitudinal data analysed in the present study are available at https://doi.org/10.57745/FHQR9Z.

The inclusion visit was performed by a gynaecologist or a midwife at the CeGIDD outside operating hours. After an interview, several samples were collected, including vaginal swabs with eSwabs (Coppan) in Amies preservation medium from which microbiota barcoding was later performed. The samples were aliquoted right after the visit and stored at –20°C, before being transferred to –70°C within a month. The participants also filled in a detailed questionnaire, which formed the basis of epidemiological covariates analysed in this study.

Subsequent on-site visits were scheduled every two or four months, depending on the HPV status. In between on-site visits, women were asked to perform eight self-samples at home with eSwabs in Amies medium and to keep them in their freezer. The self-samples were brought back in an isotherm bag at the next visit. These were then stored with the swab at –70°C until processing.

### Microbiota metabarcoding and quantification

The microbiota metabarcoding was performed on 200*µ*L of vaginal swabs specimen stored at –70*^◦^* in Amies medium. The DNA extraction was performed using the MagAttract PowerMicrobiome DNA/RNA kit (Qiagen). Next-generation sequencing of the V3-V4 region of the 16S gene [18] was performed on an Illumina HiSeq 4000 platform (150 base pairs paired-end mode) at the Genomic Resource Center at the University of Maryland School of Medicine.

The taxonomic assignment was performed using the software package SpeciateIT (https://github.com/Ravel-Laboratory/speciateIT) and the CSTs were determined using the VALENCIA software package [9]. To examine longitudinal patterns, the present study included participants who contributed at least three samples: 125 women met the inclusion criterion, giving 2,103 samples in total.

### Covariates

In the PAPCLEAR study, a questionnaire was given to each participant to record patient-level meta-data. We initially considered the following covariates based on previously proposed roles in influencing the vaginal milieu:

*1st menstr.* Number of years since the first menstruation: The morphology of the human vagina changes throughout life and the onset of puberty marks a key event that triggers cascading changes [19].

*Alcohol* Average number of glasses of alcoholic drinks consumed per week: Chronic presence of alcohol in the genital environment has been linked to a shift in the immune and microbiological conditions [20].

*Antibio.* Application of antibiotics during the study, either systemic (*Antibio. (Systemic)*) or genital (*Antibio. (Genital)*): The bacterial composition responds rapidly and transiently to antibiotic treatments that target bacteria either broadly or with a narrow taxonomic scale [21].

*BMI* Body mass index (BMI): Obesity has been implicated in elevating vaginal microbiota diversity and promoting *Prevotella* associated with BV [22].

*Caucasian* Identity as Caucasian ethnicity or other: Ethnicity has been linked to variation in vaginal microbiota compositions in several studies [6]. However, causal mechanisms remain an open question.

*Cigarettes* Cigarette smoking: Smoking has been implicated in the development of BV due to its anti-estrogenic effects and the presence of harmful substances such as benzo[a]pyrene diol epoxide (BPDE) [23].

*Horm. contra.* Use of hormonal contraception during the study: The vaginal hormonal landscape is affected by the use of hormonal contraceptives [24].

*Lubricant* Use of lubricant during the study: Personal lubricants contain various chemicals that differentially impact the growth of vaginal microbes in-vitro [25].

*Menstr. cup* Use of menstrual cups during the study: The vaginal microenvironment may be altered by the use of menstrual cups both physically and chemically. An elevated risk of fungal infections has been reported [26].

*Partners* Cumulative number of sexual partners: The genital microbiome can be transferred between sexual partners [27]. Such an external input could destabilise the resident community.

*Red meat* Average number of meals that include red meat consumption per week: Diet alters the vaginal environment for microbes. An unhealthy diet, linked to a high proportion of red meat consumption, has been linked to an elevated risk of BV [28].

*Regular condom* Regular use of condoms during sexual intercourse: Condom use can modify the vaginal microenvironment by altering the exchange of microbes between partners [29].

*Regular sport* Engaging in regular sporting activities, over 50% of the time: Physical activities influence immune responses, with leisure-time physical activity associated with a reduced risk of suspected bacterial infections compared to sedentary behaviour [30].

*Stress* Average stress level reported from 0 (min) to 3 (max): Stress hormones may disrupt vaginal flora, for instance, by inhibiting glycogen production, which is the primary fuel for lactobacilli [31].

*Tampon* Use of tampons during the study: The use of internal menstrual health products like tampons directly alters the vaginal environment, although negative effects from tampon use are seldom reported [32].

*Vag. product* Use of vaginal cream/tablet/capsule/gel/wipe during the study: Women frequently use over-the-counter vulvovaginal treatments that contain a variety of chemical components. However, the clinical effectiveness of these products in preventing BV is seldom systematically evaluated [33].

*Chlamydia* Tested positive for chlamydia.

*Female/male affinity* Affinity to female/male partner: Genital microbiome transfers during sexual activity are anticipated to vary based on the genders of the partners [34].

*Pregnancy* History of pregnancy: Pregnancy significantly changes the cervicovaginal environment, with increased estrogen from the ovaries and placenta leading to higher vaginal glycogen. This supports the growth of *Lactobacillus* species [35].

*Spermicide* Use of spermicide during the study: Spermicides are chemicals that prevent sperm from reaching an egg, but their use can change the vaginal microflora, potentially increasing the risk of genitourinary infections [36].

*Vag. douching* Use of vaginal douching during study: Vaginal douching, the practice of washing inside the vagina with a liquid solution, has been shown to increase the risk of disturbing the natural balance of vaginal flora [37].

Out of the covariates initially considered above, we excluded six (*Chlamydia*, *Female affinity*, *Male affinity*, *Pregnancy*, *Spermicide* and *Vag. douching*) as data were severely skewed towards the most common value (*>* 90% of data). During the study, any use of antibiotics was recorded with the date and we distinguished systemic (*Antibio. (Systemic)*) and genital topical (*Antibio. (Genital)*) applications, corresponding to ‘Gynecological antiinfectives and antiseptics’ (‘G01’ ATC codes), which consisted of metronidazole treatments, and ‘Antibacterials for systemic use’ (‘J01’ ATC codes), which were more diverse. Since the exact dates of treatment were recorded, *Antibio. (Systemic)* and *Antibio. (Genital)* were included as time-inhomogenous covariates in the model. All other covariates were considered time-homogeneous meaning that the variation is among women, and static through time because the precise timing of changes in the covariate values was unknown. To facilitate the comparison of covariate effects, we centred and scaled continuous variables [38] and deviation-coded binary variables. These transformations ensure that all covariates are modelled in a comparable scale and the intercept is located at a “representative reference value” of the modelled population: i.e., the population mean for continuous and the theoretical mid-point for binary values. Four continuous covariates (i.e., *Alcohol*, *BMI*, *Partners*, and *Red meat*) were log-transformed before scaling due to their right-skewed distribution. We found no strong correlations among the covariates included in the analysis (Supplementary Information S1).

## Modelling

### Markov model

Markov models are statistical models used to represent systems that transition between discrete states over time. These models are ‘memoryless’, meaning that the probability of transition to another state depends on the current state, but not its historical path. In clinical research, these models are often used to predict the transitions among health states (e.g., health, illness and remission), and the propensity to transition between these states is estimated from longitudinal follow-up data. Clinical follow-up data are typically modelled using the continuous-time Markov model [39], in which the probability of transition over a given interval depends on the instantaneous transition intensity and the amount of time spent in the current state.

Vaginal microbiota state transitions are classically studied using continuous-time Markov models [13–15, 40, 41]. Our application of the continuous-time Markov model differs from those of the existing literature in its hierarchical Bayesian formulation, which allowed us to quantify individual variability among women (as unobserved heterogeneity, or random effects) and to estimate many covariate effects simultaneously (through the use of weakly informative priors).

### Transition intensities

Transition intensities, *q*, refer to the instantaneous rate of moving from state *i* to state *j* in a participant *p* (e.g., CST I to CST IV), a process that may be affected by a vector of covariates, *X*. Taking the form of a proportional hazards model, these rates can be expressed as:

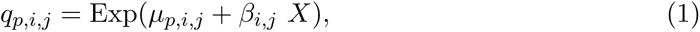

where *µ_p,i,j_* is the intercept and *β_i,j_* is the coefficient expressing the impact of a covariate(s). This intercept is further defined by the equation,

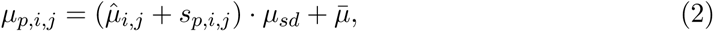

where 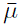 and *µ_sd_* are the prior mean and standard deviation of the intercept such that 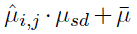 *s*constitutes the non-centred parameterisation of the population-level intercept, *µ_i,j_* and is assumed to be normally distributed, i.e., 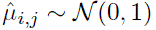.

Additionally, we allowed for unobserved heterogeneity in *µ*, i.e., *s_p,i,j_*, where

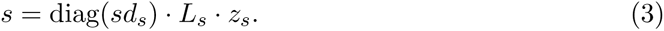

We sampled from the corresponding weakly informative priors, namely *sd_s_* ∼ *t*_4_(0, 1), *L_s_* ∼ LKJCorrCholesky(2) (which slightly favours correlations among unobserved heterogeneity closer to zero, reducing the likelihood of extreme positive or negative correlations), and *z_s_*∼ N (0, 1), as recommended by the Stan development community [42, 43]. The multivariate normal density and the LKJ prior require the matrix parameters to be decomposed, which can be computationally intensive if done repeatedly. To ensure computational efficiency and numerical stability, the model was directly parameterised using the Cholesky factors of correlation matrices. This approach uses a multivariate version of the non-centred parameterisation.

For regression coefficients, the Student-t distributions with degrees of freedom 4 to 7 are recommended as generic, weakly informative, priors [43]: we sampled *β* from *β* ∼ *t*_4_(0, 1), which places a comparatively wide tail within the recommendation. As all of our covariates have been proposed to impact vaginal microbiota communities *a priori* (see above), we did not strongly regularise the priors, for example, through the use of horseshoe priors [44].

We note that all covariates were modelled simultaneously, such that the interpretation of each coefficient is conditional upon other covariates included and accounts for the influence of other factors. We assumed that the covariates affect the transitions symmetrically (i.e., *β_j,i_* = −*β_i,j_*), meaning that the influence of a covariate on the affinity (or aversion) towards a particular CST is consistent, regardless of the direction of the transition.

Collectively, the transition intensities form the matrix, *Q_p_*, in which the sum of intensities across a row, i.e., all transitions from a particular state, is defined to be zero, such that we have the following equation for the diagonal entries [39]:

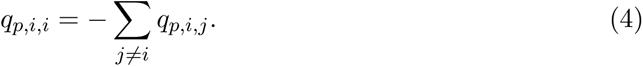

### Transition probabilities and likelihood

Taking the matrix exponential of the *Q_p_* matrix for each participant, *p*, we compute the matrix *P_p_* such that:

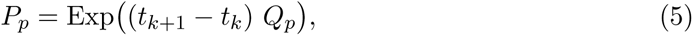

where *k* represents the sample identity for a given individual. The *P_p_* matrix contains the transition probabilities between two observations (at *k* and *k* +1) and *t_k_*_+1_ −*t_k_*indicates the elapsed time between two observations.

Finally, the probability of observing a given state at the next sampling event (i.e., at *k* + 1) is modelled by the categorical distribution:

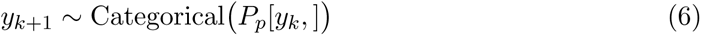

where *P_p_*[*y_k_,*] is the *y_k_*^th^ row of the *P_p_*matrix containing the probabilities of transition from the state observed at *k*.

### Model fitting

We used a Bayesian approach to fit the above continuous-time Markov model to longitudinal data of vaginal microbiota CSTs. In total, the model consisted of 57 parameters and 12 hyper-parameters. Our model was written in Stan 2.26.1 and fitted through the RStan interface 2.32.3 [45]. The Stan programme is available at https://doi.org/10.57745/FHQR9Z. One participant lacked information on the years since their initial menstruation. We imputed missing values using the mice package [46] and generated 10 imputed datasets to be fitted separately. For each imputed dataset, we fitted the model in parallel using four independent chains, each with 10, 000 sampled iterations and 1, 000 warm-up iterations. The MCMC samples from separate runs (i.e., based on differently imputed data) were subsequently combined for inference.

We confirmed over 1, 000 effective samples per imputed dataset and ensured convergence of independent chains (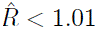) for all parameters [47]. We carried out a posterior predictive check by comparing the observed and predicted CST frequency. We also quantified the posterior *z*-score and posterior contraction to examine the accuracy and precision of posterior distributions and the relative strength of data to prior information [48] (Supplementary Information S2).

### Counterfactual predictions

We took advantage of the parameterised model to simulate the population-level outcomes of each covariate, assuming that all covariates, but a focal one, are at the representative reference value (as described above) and then varying the focal parameter within the range of values observed in the studied cohort. The model predictions were generated by randomly drawing 100 samples from the posterior distributions and simulating the Markov model for each sampled parameter set. We focused on the CST frequency as the outcome of interest.

## Results and Discussion

### CSTs in the cohort

As is typical of vaginal microbiota communities, the microbial compositions sampled in PAPCLEAR were highly structured, and characterised by a relatively small number of operational taxonomic units (OTUs). The dominant species within these communities aligned closely with specific community state types (CSTs) as defined by Ravel et al. [6]. For example, CST I was primarily associated with *L. crispatus* and CST III with *L. iners*.

In contrast, and as expected, CST IV communities exhibited a higher degree of microbial diversity compared to CSTs dominated by lactobacilli, reflecting a broader range of species typical of this community type (Fig. 1).

**Figure 1:**
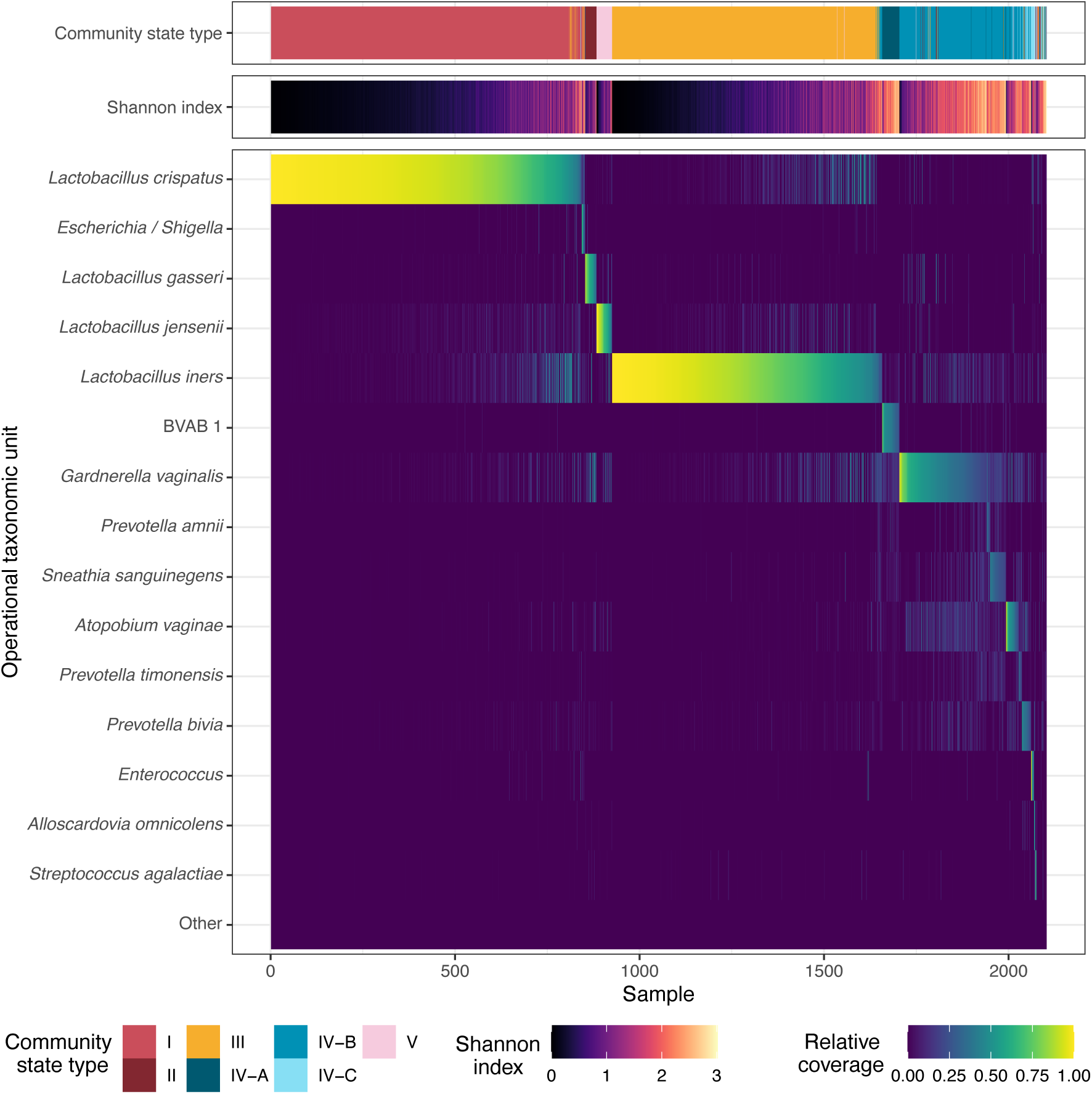
Vaginal community state types (CSTs), diversity (Shannon Index), and relative coverage of the 15 most common taxonomic operational units (OTU) of 2,103 samples from the PAPCEAR cohort. In over 98.5% of samples, a single of these 15 OTUs represented the most common OTU.

Our longitudinal dataset from the PAPCLEAR cohort represents one of the largest analysed to date in the context of the vaginal microbiota. Detailed participant characteristics are presented in Table 1. Briefly, the participants were between 18 and 25 years old and the majority of the 2,103 samples (73.7%) were self-collected at home, the rest being collected during on-site visits (Fig. 2a). The median follow-up duration was 8.64 months and the most common intervals between analysed samples were seven and 28 days (Fig. 2a & b). On average, each of the 125 participants contributed 11 samples (Fig. 2c).

**Figure 2:**
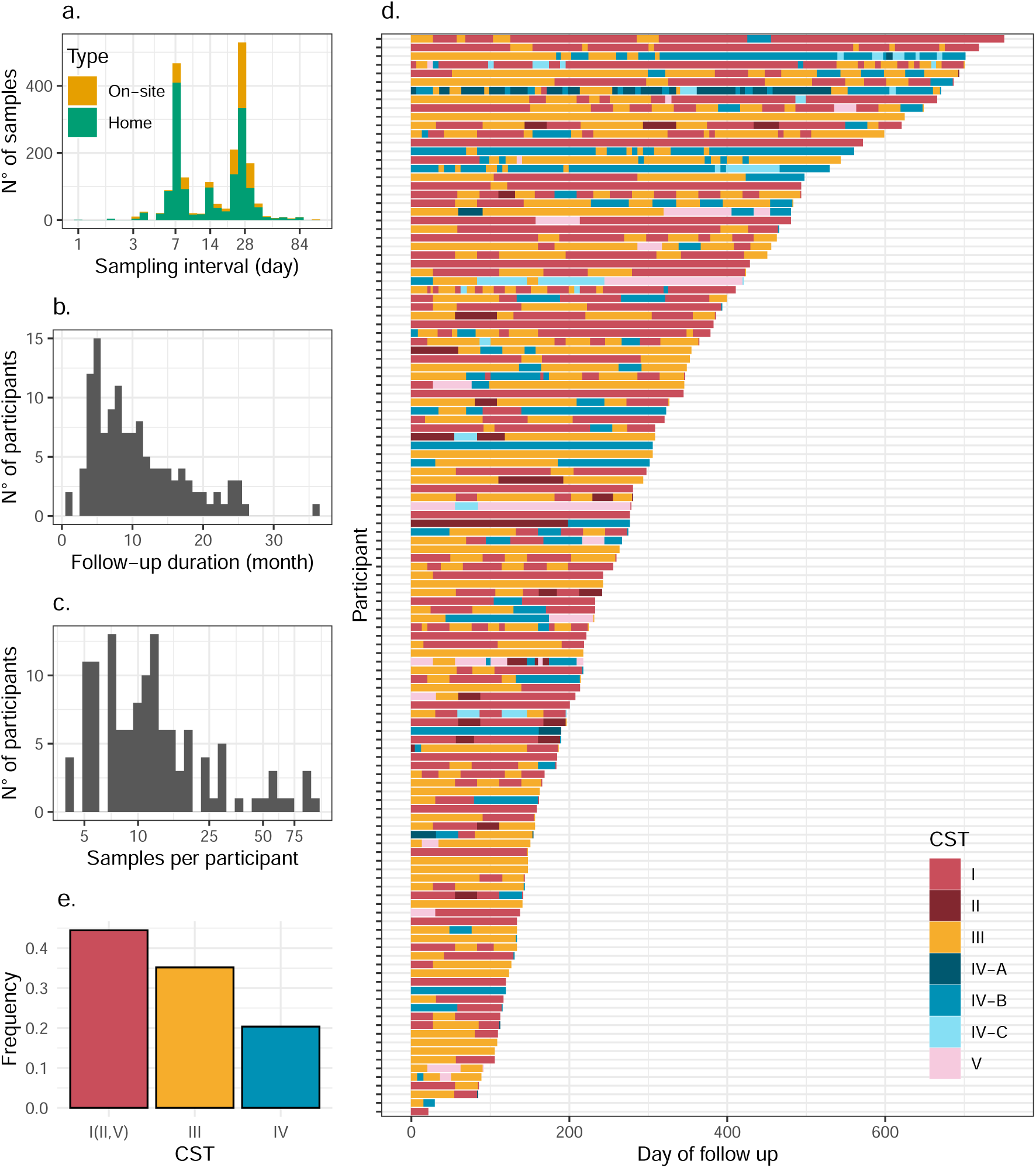
Summary of vaginal microbiota samples analysed in the PAPCLEAR study. a) Intervals between sampling events for clinical (i.e., on-site) and home samples. b) Follow-up duration per participant. c) Number of samples analysed per participant. d) Vaginal microbiota Community State Types (CST) over time in 125 participants. For visualisation, data are truncated at 750 days for a single individual whose duration exceeds this threshold. e) Frequency of the optimal (i.e., CSTs I, II, and V combined), sub-optimal (CST III) and non-optimal (CST IV) communities in all samples.

**Table 1:** Summary profile of vaginal microbiota samples and covariates in the PAPCLEAR study. Q1 and Q3 refer to first (25%) and third (75%) quantiles. Level = 1 indicates the presence of a binary condition. See Materials and Methods for the covariate definitions.

|  | Level | Summary |
| --- | --- | --- |
| Samples (Participants) |  | 2103 (125) |
| CST (%) | I | 847 (40.3) |
|  | II | 39 (1.85) |
|  | III | 740 (35.2) |
|  | IV-A | 54 (2.57) |
|  | IV-B | 342 (16.3) |
|  | IV-C | 32 (1.52) |
|  | V | 49 (2.33) |
| Sample type (%) | On-site | 553 (26.3) |
|  | Home | 1550 (73.7) |
| Sampling interval (median (Q1,Q3)) |  | 21 (7, 28) |
| Follow-up duration (median (Q1,Q3)) |  | 8.64 (5.36, 14.0) |
| Samples per subject (median (Q1,Q3)) |  | 11 (7, 16) |
| *Covariates* |  |  |
| Identifying as ‘Caucasian’ (%) | 1 | 102 (81.6) |
| BMI (median (Q1,Q3)) |  | 21.19 (19.78, 23.46) |
| Alcohol (median (Q1,Q3)) |  | 3.14 (1.40, 5.07) |
| Smoker (%) | 1 | 36 (28.8) |
| Stress level (from 0 to 3, median (Q1,Q3)) |  | 1.41 (1.00, 1.75) |
| Regular sport practice (%) | 1 | 61 (48.8) |
| Red meat consumption (times per week, median (Q1,Q3)) |  | 0.50 (0.16, 1.00) |
| Years since 1st menstruation (median (Q1,Q3)) |  | 9 (7, 10) |
| Hormonal contraception (%) | 1 | 32 (25.6) |
| Menstrual cup user (%) | 1 | 46 (36.8) |
| Vaginal product user (%) | 1 | 73 (58.4) |
| Tampon user (%) | 1 | 89 (71.2) |
| Lifetime number of partners (median (Q1,Q3)) |  | 5 (3, 11) |
| Lubricant use (%) | 1 | 58 (46.4) |
| Regular condom use by partner (%) | 1 | 23 (18.4) |
| Male affinity (%) | 1 | 124 (99.2) |
| Chlamydia infection at inclusion (%) | 1 | 7 (5.6) |
| Pregnancy during follow-up (%) | 1 | 4 (3.2) |
| Vaginal douching (%) | 1 | 4 (3.2) |
| Spermicide user (%) | 1 | 1 (0.8) |
| Female affinity (%) | 1 | 10 (8.0) |
| Systemic antibiotic treatment (%) | 1 | 65 (52.0) |
| Genital antibiotic treatment (%) | 1 | 30 (24.0) |

The metabarcoding analysis on 16S RNA with the VALENCIA algorithm [9] was used to assign each sample to a CST. The vaginal microbiota communities were variable across women and over time (Fig. 2d). As CSTs I, II, and V are all dominated by lactobacilli and considered ‘optimal’ in terms of health, yet the latter two are rare (∼4% of all samples combined), we pooled the three optimal communities for further investigation. Overall, optimal communities were the most frequent, representing 44.5% of samples, followed by ‘sub-optimal’ (CST III) at 35.2% and ‘non-optimal’ communities (CST IV) at 20.4% (Fig. 2e and Table 1).

### Probabilities of CST persistence

We implemented a continuous-time Markov model to capture the CST dynamics. Simulations based on the estimated parameters of our model (i.e., posterior predictive check) confirmed that it accurately captures the observed CST prevalence (Fig. 3a). The optimal, CST I (II, V), and sub-optimal, CST III, communities showed a high degree of stability, with weekly probabilities to remain in the current state estimated at 87% (95% credibility interval (95CrI): 78 – 93%) and 81% (95CrI: 68 – 90%), respectively (Fig. 3b). In contrast, the weekly persistence probability of the non-optimal CST IV was 60% (95CrI: 35 – 80%, Fig. 3b). These transition probabilities translate into sojourn times (i.e., the expected time spent in a given state before moving to another) in CST I (II, V), III and IV of 6.9 days (95CrI: 2.9 – 13.6 days), 4.23 days (95CrI: 1.8 – 8.4 days) and 1.6 days (95CrI: 0.58 – 3.8 days), respectively.

**Figure 3:**
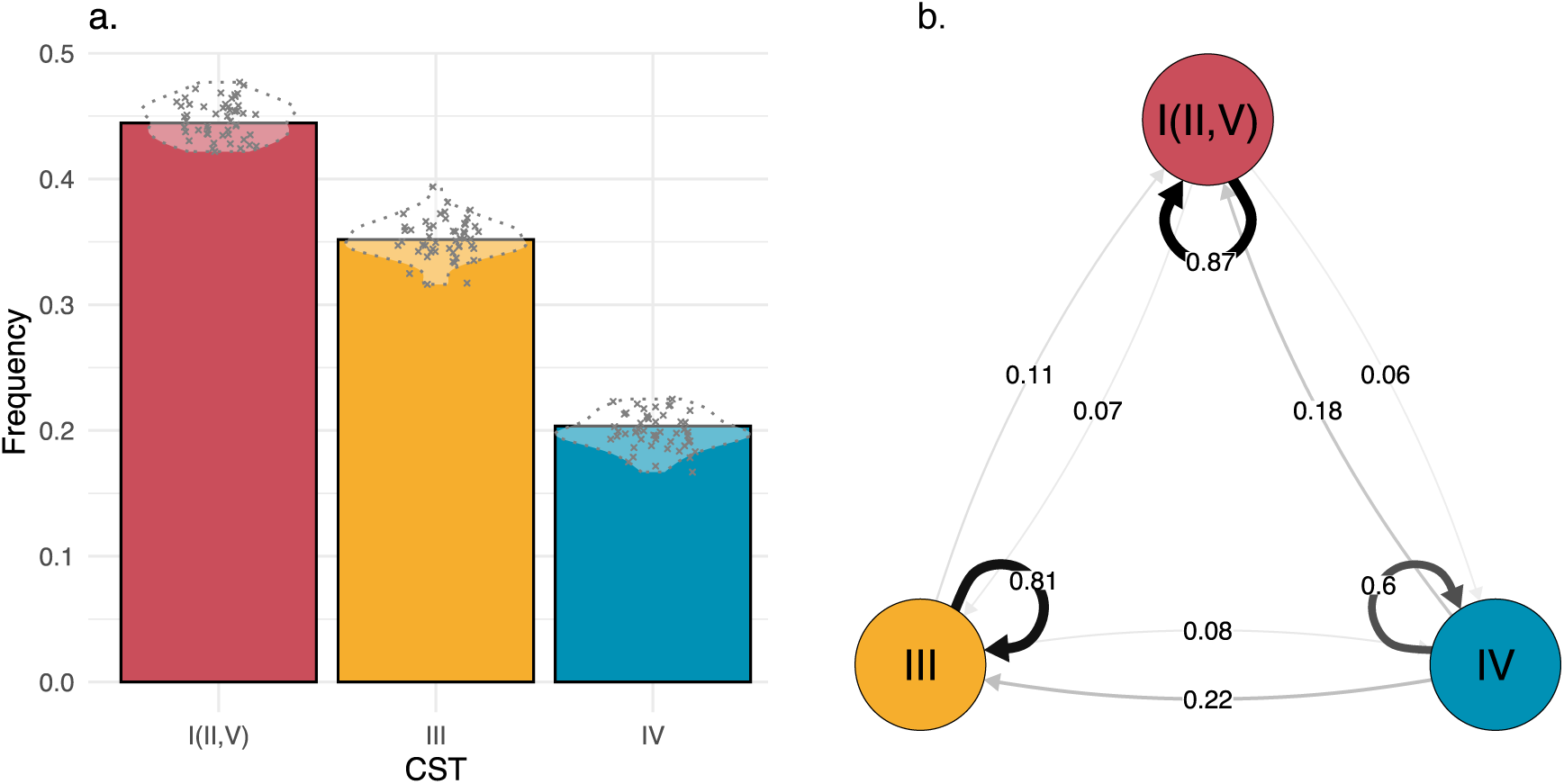
Prevalence and transition probabilities among vaginal microbiota community state types (CSTs). a) Observed (bars) and predicted prevalence (crosses) of CSTs I (II, V), III and IV. The model predictions were generated by drawing 100 random samples from the posterior distributions and simulating the Markov model for each sampled parameter set. b) Mean estimated weekly transition probabilities of CSTs I (II, V), III and IV. The arrow thickness indicates the persistence or transition probability.

The reported persistence and transition probabilities in the literature vary widely based on the cohort characteristics. For example, focusing on women during pregnancy, DiGiulio et al. [13] estimated that the four *Lactobacillus*-dominated CSTs (CSTs I, II, III, and V) were more stable than CST IV. Notably, both CST I and II showed 98% probability of weekly persistence. The enhanced persistence of *Lactobacillus*-dominated communities during pregnancy owes itself to specific vaginal conditions during pregnancy including the up-regulation of oestrogen and progesterone that facilitates lactobacilli [13, 49].

In addition, the temporal dynamics of vaginal microbiota are notably different in women with BV. In contrast to pregnant women, those experiencing symptomatic BV generally exhibit less stable vaginal microbiota communities. In the cohort of Ravel et al. [50], which focused on women with symptomatic BV, Brooks et al. [40] found significantly lower stability across all CSTs. The probability of these CSTs persisting ranged from 38% to 48%, with CST I persisting only 46% of the time over a week.

Among studies that focused on non-pregnant, healthy young women — with no particular emphasis on BV — the analysis by Brooks et al. [40] of the Chaban et al. cohort [16] (N = 27; Canada) estimated weekly persistence probabilities of 75% for CST I, 78% for III, 60% for IV-A, and 88% for V. In the Gajer et al. dataset [12] (N = 32; USA), analysed again by Brooks et al., [40], CST I, II and III demonstrated 72%, 84% and 77% weekly persistence probabilities, respectively. In this dataset, CST IV sub-categories showed markedly different stability with CST IV-A with weekly persistence of 38% and CST IV-B with persistence of 82%. A third study, Munoz et al. [15] (N = 88; South Africa), reported the stability of vaginal microbiota in women in a three-month time frame using a different microbiota classification system consisting of four categories predominantly associated with: *L. crispatus* (similar to CST I), *L. iners* (similar to CST III), *G. vaginalis* (similar to CST IV), or *Prevotella* spp. (similar to CST IV). They found similar persistence for CST I and CST IV-like communities ranging from 51 to 53% over three months while the CST III-like community was more stable at 62% over the same period. Recasting in the three-month time scale, our estimates show the same extent of stability for CST I(II, V) at 51% (95% CrI: 29-72%) while CST III (38%, 95% CrI: 19-61%) and CST IV (15%, 95% CrI: 5-34%) were less stable. Taken together, our estimates of vaginal microbiota community stability are within the range of values reported in other cohorts. However, the dynamics of vaginal microbiota communities are likely geographically variable even among healthy young women.

### Covariate effects on transitions

The Bayesian approach, which can accommodate vaguely informative priors on the covariate effects, allows for the simultaneous inclusion of many covariates (as hazard ratios; Eq. 1) which would otherwise prove difficult in Markov models [39]. We identified 16 covariates based on previously proposed roles in influencing the vaginal milieu and assumed that covariates have a symmetrical effect on CST transitions: e.g., the magnitude of a given covariate effect on the transition from CST I to III is identical to that on the transition from CST III to I. We identified alcohol consumption as the strongest and most consistent effect while several other covariates were identified as possible drivers of CST transitions.

### Alcohol consumption

The estimated hazard ratios on community transitions indicate that alcohol consumption favoured the sub-optimal (CST III) community over optimal (CST I(II, V)) with 97% probability (Fig. 4). Because of our symmetry assumption, this can mean that alcohol consumption increases the pace of transition from CST I(II, V) to CST III or reduces that in the opposite direction by the same magnitude. Alcohol consumption also tended to favour CST IV over CST III, although with a lower credibility level (with 73% probability of the hazard ratio ≠ 1, Fig. 4).

**Figure 4:**
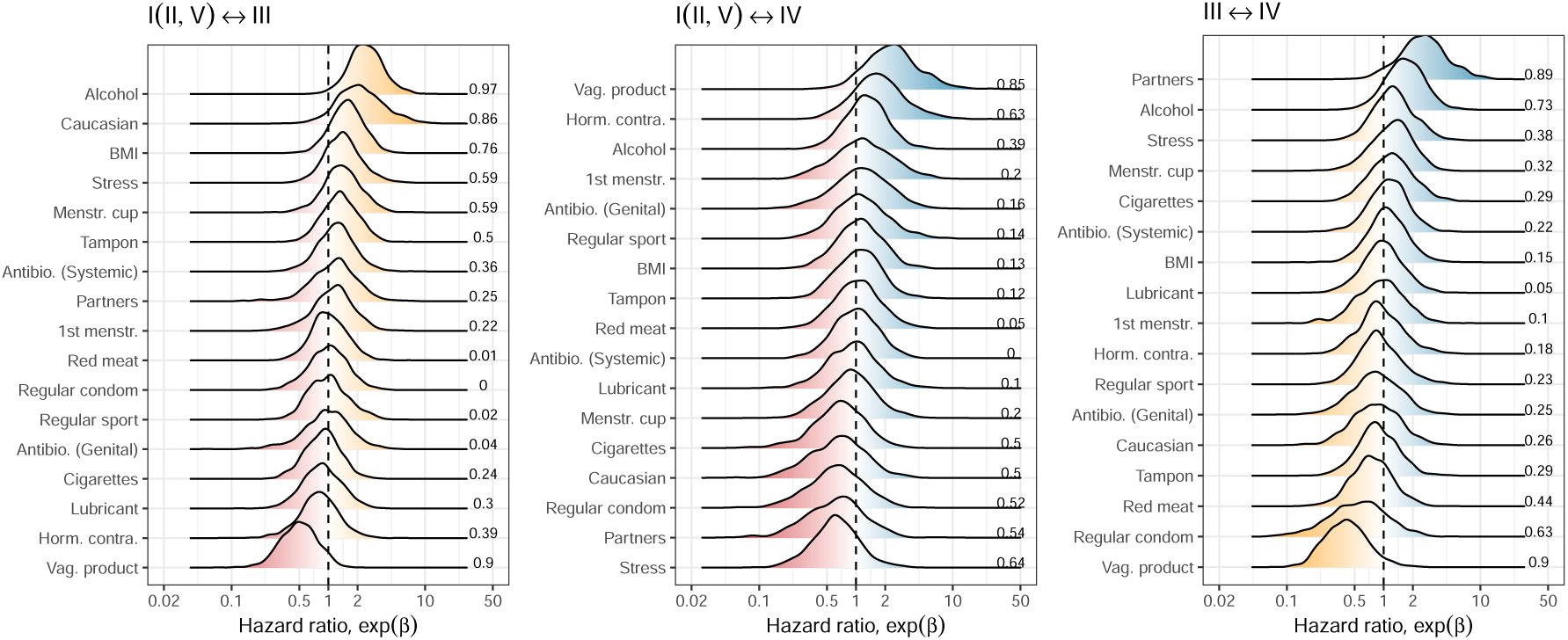
Estimated covariate effects on community transition rates. With the symmetry assumption, there are three sets of covariate effects on transitions. The impact of covariates on community transition rates was estimated for a given set of community states as the log hazard ratio, *β*. The figure shows the posterior distributions of exp(*β*), the hazard ratio for the three sets of transition sets, and the corresponding 16 covariates. The numbers on the right-hand side of each panel indicate the probability that the estimated effect is different from the hazard ratio of 1 (i.e., the proportion of posterior distributions sampled on the dominant side of the effect). For example, alcohol consumption was estimated to favour CST III over CST I (II,V) at a credibility level of 97%.

To examine how these effects translate to the population level, we carried out counterfactual simulations in which all participant characteristics were set to the representative value observed in the studied cohort, except for alcohol consumption, which ranged from non-drinking to the level of the heaviest drinking observed in our cohort (19 drinks per week). The simulations demonstrated that the expected prevalence of the optimal (CST I (II, V)) community was 18% (95% CrI of 9 to 27%) higher in a hypothetical population of non-drinkers compared to that of average-level drinkers who consumed three drinks per week (Fig. 5; Supplementary Information S3). In turn, the prevalence of the optimal community was 19% (95% CrI of 10 to 29%) higher in the population of average-level drinkers than in the heaviest drinkers. As the optimal community declined with alcohol consumption, the prevalence of the non-optimal (CST IV) community was found to be 9% (95% CrI of 2 to 15%) higher among average drinkers compared to non-drinkers. Therefore, while the strongest impact of alcohol on community transitions appears to be between the optimal (CST I (II, V)) and sub-optimal (CST III) communities, an additional, non-zero impact on the sub-optimal to non-optimal (CST IV) transition means that alcohol consumption ultimately promotes non-optimal communities at the expense of optimal ones. As the effects of covariates are estimated simultaneously, potential confounding factors, including the number of partners, condom use and smoking, are controlled for in our findings.

**Figure 5:**
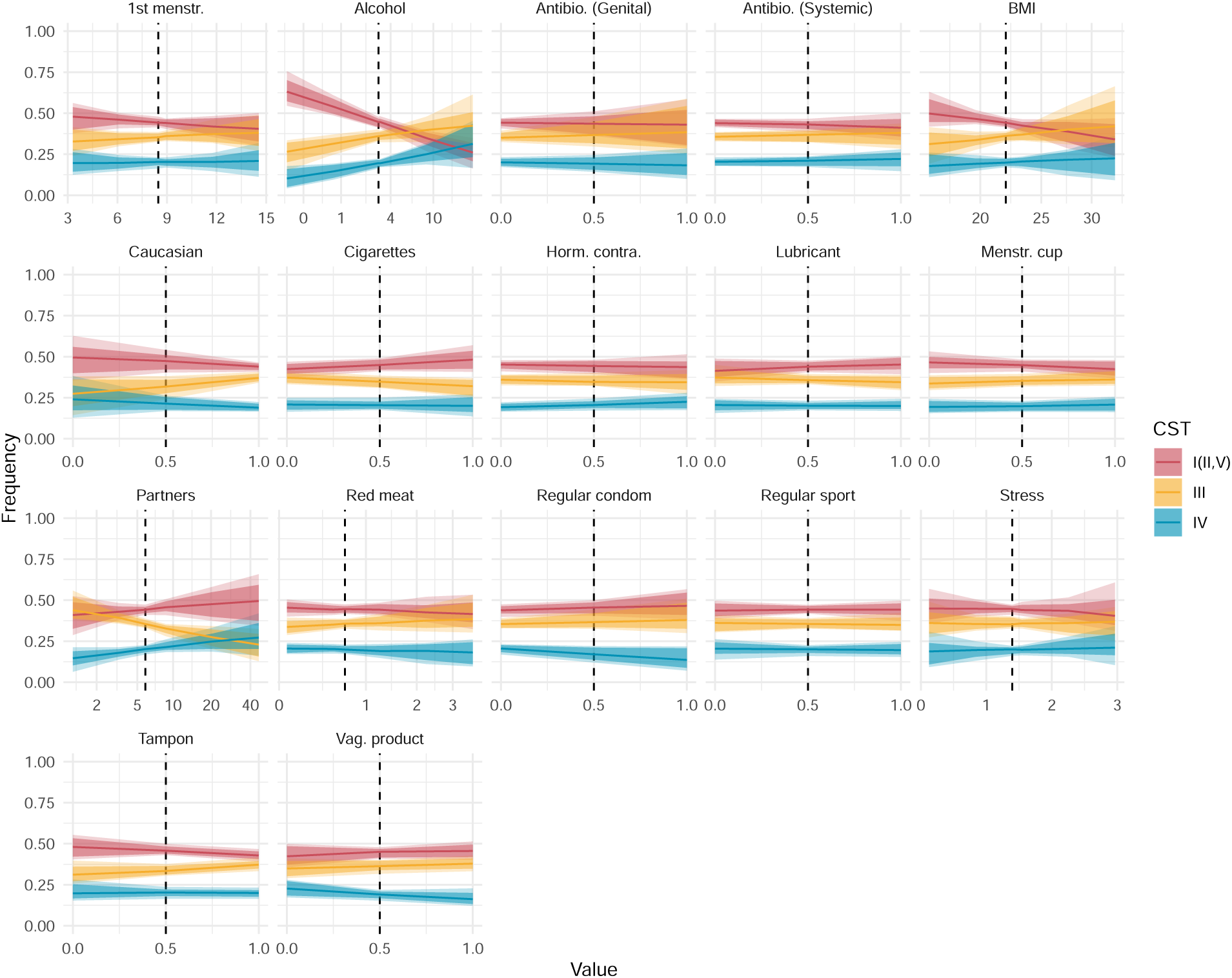
Counter-factual simulations predict population-level consequences of covariates. Based on estimated hazard ratios (Fig. 4), the population-level impact (i.e., the prevalence of CST I (II,V), III and IV) was simulated for each covariate. The vertical dashed lines indicate the intercept used in estimation: i.e., the population mean for continuous and midpoint for binary variables. For continuous variables, the range of values explored was determined by the minimum and maximum values reported in the PAPCLEAR study.

Alcohol consumption may impact the vaginal microbiota through a variety of mechanisms. Physiologically, the chronic presence of alcohol in the genital environment has been linked to a shift in immune and microbiological conditions [20]. In addition, alcohol is a known modifier of sexual behaviour, which in turn has been demonstrated to increase the risk of BV, linked to CST IV [51]. Finally, alcohol alters the microbial profile in other body parts, which in turn could cross over to the vaginal milieu. For example, *Prevotella*, a genus commonly found in CST IV communities, is enriched in the oral microbiota of drinkers [52]. Similarly, others postulate the effect of alcohol on the gut microbiota may have a concurrent influence on the vaginal microbiota [53].

While there remains a lack of consensus among existing studies (briefly reviewed by Froehle et al. [53]), cohort and cross-sectional studies from diverse geographical contexts (namely, Australia, Denmark, Sweden, Thailand, Tanzania, Uganda and USA) have previously reported an association between alcohol consumption and BV [53–61]. In addition to corroborating these findings, our Markov model offers a novel insight into the ecology of microbial communities underlying these observations: alcohol consumption destabilises the optimal (CST I (II, V)) communities towards sub-optimal (CST III), which opens the gate for the deterioration towards non-optimal (CST IV), associated with BV. To the authors’ knowledge, there have been no alcohol cessation studies reporting its impact on vaginal microbiota. Such studies are necessary to establish causal links, similar to those conducted on the effects of smoking [23], douching [62], and antibiotics [21] on vaginal microbiota compositions.

### Potential effects of other covariates

Other factors with possible effects on transitions (i.e., with more than 80% probability of hazard ratio ≠ 1) included the use of vaginal intimate hygiene products, number of sexual partners and self-reported ‘Caucasian’ identity.

#### Vaginal hygiene products

The use of vaginal hygiene products, defined broadly here to include vaginal cream, tablet, capsule, gel and wipe, appeared to have multifaceted effects. Between CST I (II, V) and CST III, their use was positively linked to maintaining or transitioning to CST I (II, V) with 90% probability (Fig. 4). For the CST I (II, V) and CST IV pair, it tended to favour a shift towards CST IV, with 85% probability. Finally, between CST III and CST IV, their use was more likely to support the persistence or a shift towards CST III, also with 90% probability. The circular effects suggest that women may experience different effects of the products marketed for ‘vaginal intimate hygiene’ depending on the predisposition with certain CSTs. Nonetheless, the circular effects on community transitions meant that there was no noticeable impact at the population level in our counterfactual simulations (Fig. 5).

#### Number of sexual partners

A higher number of sexual partners was also found to potentially favour CST IV over CST III, increasing the risk of maintaining (or transitioning to) CST IV with 89% probability of the hazard ratio ̸= 1. The association between CST IV and the lifetime number of partners is consistent with the hypothesis that external importation of microbes could alter the dynamics of vaginal microbiota and is in line with earlier work [63, 64]. Population-level simulations predict that an increasing number of sexual partners tends to reduce the prevalence of the sub-optimal (CST III) community. For example, CST III was 13% (95% CrI of 2 to 21%) less common in a hypothetical population with the highest number of partners than one conforming to the average number. The decrease was accompanied by a tendency for the other CSTs to increase, although the trend was less clear for CST I(II,V) and CST IV, individually (Fig. 5).

#### Causasian identity

It is worth noting that our cohort was not designed to achieve comprehensive coverage of self-reported ethnic identity, with over 80% identifying as Caucasians (Table 1). Nonetheless, identifying oneself as a ‘Caucasian’ tended to favour CST III over CST I(II, V) with 86% probability. European studies focusing on the role of ethnicity are rare. However, a North American study has observed a qualitatively opposite trend: CST III communities are comparably rare in women who identify themselves as Caucasian compared to those identifying as Asian, Black and Hispanic (26.8 versus 42.7, 31.4 and 36.1%, respectively [6]). While previous studies have revealed differences in vaginal microbiota compositions among ethnic groups, the relative importance of biological, societal, and environmental factors remains an open question [6, 65–67].

#### Antibiotics

Notably, we found little association between antibiotic consumption and CST transitions, neither for local treatment for BV (genital application of metronidazole) nor systemic treatment (antibiotic treatment via oral intake). Such a lack of effect in our study may be because the changes in the vaginal microbiota compositions following an antibiotic treatment take place in a shorter time scale than our sampling intervals: the most common sampling intervals were either 7 or 28 days (Table 1). In comparison, Brooks et al. [40] found rapid CST transition following BV medication in the cohort of Ravel et al. [50], which involved daily sampling. On a longer time scale, the re-emergence of BV-associated communities following treatments is a well-documented clinical challenge [68–70].

### Unobserved individual variability in community transition

While we incorporated 16 covariates into our Markov model, some variations among women remain unaccounted for. To quantify these, we estimated the extent of individual variability (i.e., unobserved heterogeneity, or random effects) in community transitions for each transition pair using a hierarchical Bayesian approach (Eq.2 & 3).

The highest variability was observed among women in the transitions involving ‘recovery’ to an optimal (CST I (II,V)) from a non-optimal (CST IV) state (Fig. 6). On the other hand, inverse transitions from optimal to non-optimal exhibited some of the lowest individual variability. The same is true, although to a lesser extent, for the shifts from sub-optimal (CST III) to optimal. These findings suggest that there are relatively limited pathways leading to the deterioration of vaginal microbiota communities, whereas the routes to recovery can be more individualised and the source of this variation remains to be fully elucidated.

**Figure 6:**
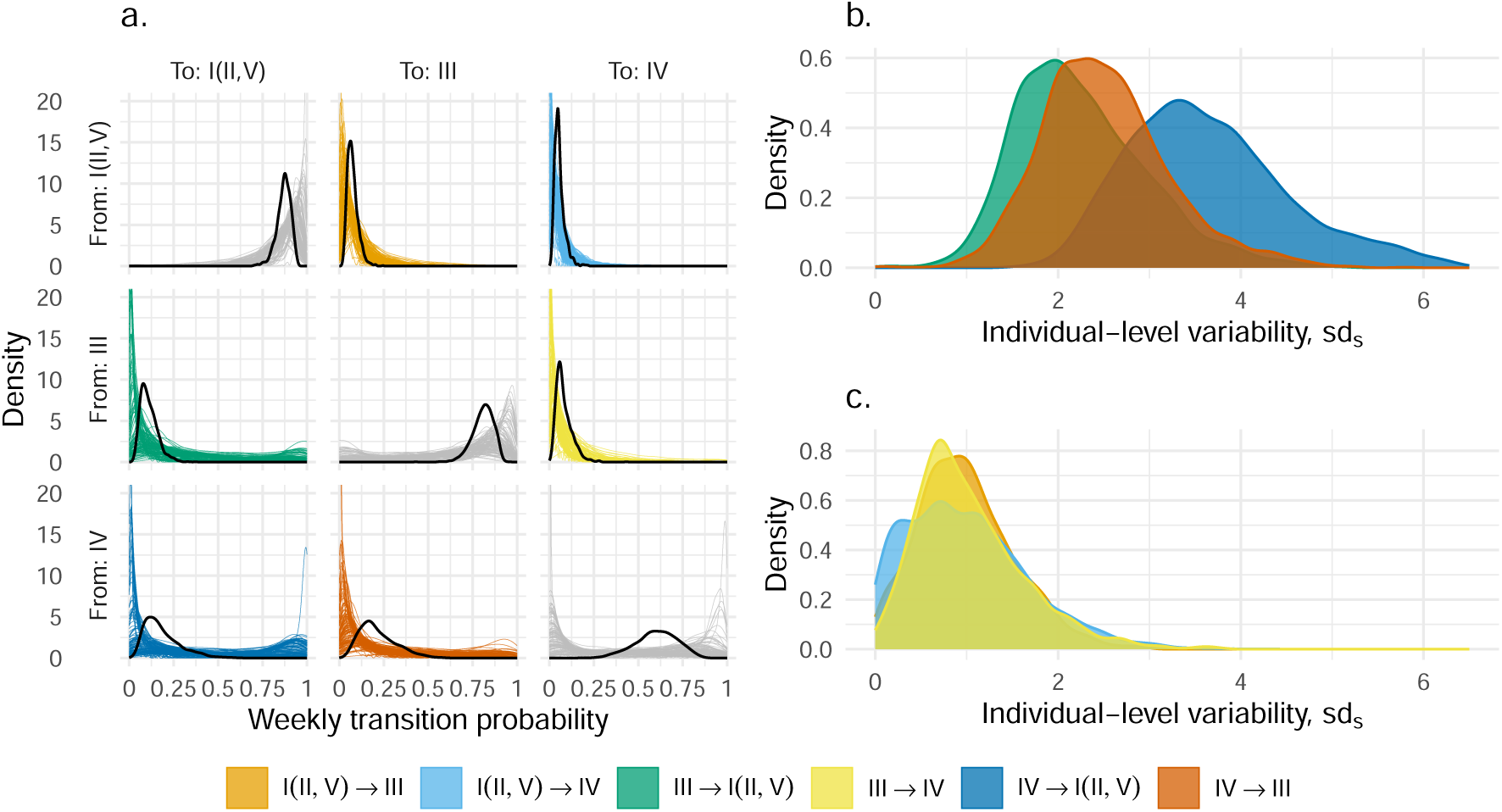
Individual-level variability in vaginal microbiota community state type (CST) transitions. a) The population average (thick black) and individual (thinner colours) weekly transition probabilities. Between-women individual variation for transitions to b) a more optimal and c) a less optimal state. Colours indicate the type of transition between CSTs.

The presence of individual-level random effects indicates that a considerable part of the variability remains unaccounted for by the 16 covariates in this study. One possible cause is that our study left out key drivers of the vaginal milieu. For example, while menstrual cycles have been demonstrated to influence daily and weekly transitions [12], they were omitted from our analysis because the timing of menstruation was ambiguous in the PAPCLEAR study. Furthermore, while large-scale longitudinal studies present logistical challenges, a citizen-science-based approach offers the potential for expanding the cohort size, thereby enhancing the statistical power needed to examine additional covariates [71]. Secondly, further resolution on individual variability may be gained by incorporating time-varying covariates, which could accommodate changes in participant behaviours during the followup. In continuous-time Markov models, time-varying covariates are assumed piecewise constant, meaning they are constant between sampling events [39]. Such an assumption is convenient as covariate values are rarely known between sampling events. Without precise knowledge of the timing of the covariate changes, however, it is unclear whether the previous covariate value (at *t* − 1) or the new covariate value (at *t*) should influence the transition.

Consequently, our analysis focused on static covariates, with the exception of antibiotic treatments for which the exact application dates were known. Aggregating participant behaviours as static covariates eliminated the uncertainty of covariate dynamics, albeit at a potentially lost opportunity for further precision.

### Limitations and opportunities

A potential limitation from a clinical methodological perspective is that the majority of samples were collected at home during the PAPCLEAR study. While home sampling could introduce variability, the participants were provided with detailed instructions to minimise the difference in swabbing techniques between on-site and home samples, and we verified consistency in sampling dates by having participants fill out online questionnaires during sampling.

Another possible limitation of our study is the resolution of microbiota community classification. We focused on three CST groups with varying health implications: optimal (CST I (II,V)), sub-optimal (III), and non-optimal (IV). This decision stemmed from the fact that detailed classifications in a Markov model would increase the number of possible transitions, and it would be difficult to estimate transitions between rare types. However, significant functional differences may exist within these CSTs. For instance, the VALENCIA algorithm classifies subcategories within some CSTs [9], and Brooks et al. demonstrated that CST IV-B is more stable than CST IV-A [40]. We also note that there are several clustering algorithms of microbial communities besides the CST framework [71, 72], which may offer differing insights on community transitions. Furthermore, the centroid distance computed by VALENCIA for CST assignment may also be leveraged to develop a quantitative, multi-dimensional perspective of the vaginal microbiota communities. Such a quantitive perspective may enhance our understanding of within-CST variabilities — although we are unaware of an existing approach that accommodates the temporal patterns in such data. Finally, the metagenomics approach holds the promise to uncover within-species diversities: e.g., metagenomics CSTs (MgCSTs) have identified with 25 distinct communities [73]. Such an approach helps to identify lineage replacements in women with stable CSTs and investigating the impact of antibiotic treatments on the prevalence of resistance genes could yield insights into the within-species dynamics of vaginal microbes. A promising direction for future research is the joint analysis of CST dynamics and sexually transmitted infections such as HPV. Previous studies have found a weak association between CST IV and HPV detection risk [74]. However, these studies tested the CST effect after estimating transition rates and pooled all high-risk and low-risk HPVs, making it difficult to identify coinfections or reinfections. The PAPCLEAR cohort, with genotype-specific follow-ups, could provide new insights into the link between CST and HPV infection, potentially identifying causal relationships.

## Conclusion

We showcased a novel application of a hierarchical Bayesian Markov model to original clinical cohort data of vaginal microbiota dynamics. Our approach facilitated the simultaneous estimation of several covariate effects on community transitions and the identification of unobserved variability in these transitions. Our work paves the way for an improved ecological understanding of microbial dynamics within the vaginal environment and indicates lifestyle alterations (such as reduced alcohol consumption) that may promote vaginal health.

## Ethics

This study has been approved by the Comité de Protection des Personnes (CPP) Sud Méditerranée I (reference number 2016-A00712-49); by the Comité Consultatif sur le Traitement de l’Information en matière de Recherche dans le domaine de la Santé (reference number 16.504); by the Commission Nationale Informatique et Libertés (reference number MMS/ABD/ AR1612278, decision number DR-2016–488), by the Agence Nationale de Sécurité du Médicament et des Produits de Santé (reference 20160072000007), and is registered at ClinicalTrials.gov under the ID NCT02946346.

## Data Availability

All the data analysed in the present work along with the scripts are available at https://doi.org/10.57745/FHQR9Z

## Acknowledgements

The authors thank Olivier Supplisson for his helpful feedback. We acknowledge the ISO 9001 certified IRD i-Trop HPC (member of the South Green Platform) at IRD Montpellier for providing HPC resources that have contributed to the research results reported within this article (bioinfo.ird.fr and www.southgreen.fr). DNA extracts were (partly) performed through the genotyping and sequencing facilities of ISEM (Institut des Sciences de l’Evolution-Montpellier) and Labex Centre Méditerranéen Environnement Biodiversité.

## Supplementary Information

### S1: Pairwise correlations between covariates

There were no strong correlations among covariates, with the strongest correlation found between BMI and stress (*r* = 0.41; Fig. S1).

**Figure S1:**
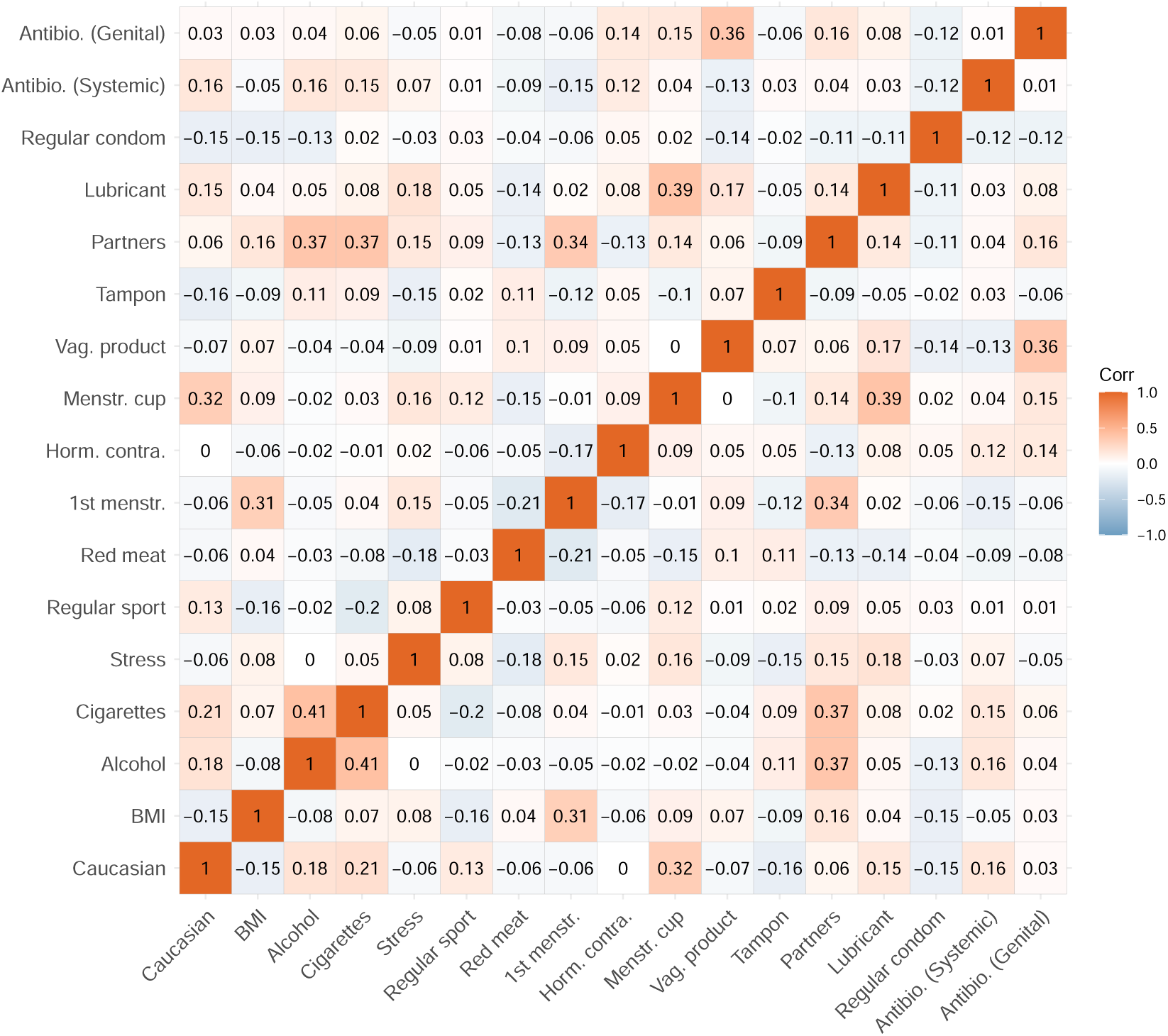
Correlation between covariates. Pairwise Pearson’s correlation coefficients between covariates. Parameter descriptions are found in Materials and Methods.

### S2: Assessment of posterior accuracy, precision and prior contraction

We leveraged the properties of posterior distributions to identify potential model fitting problems that might manifest from our model assumptions. To examine the accuracy and precision of posterior distributions, we first generated simulated observations based on the estimated posterior mean parameters. We then refitted our model to the simulated observations (i.e., secondary fitting) to compute the posterior z-score for each parameter, which measures how closely the posterior recovers the parameters of the data generating process [48]:

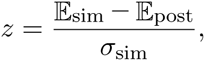

where E_post_ denotes the posterior mean of the fit to the actual data that we consider the ‘true’ parameter. E_sim_ and *σ*_sim_ denote the mean and standard deviation of the posterior distribution of the secondary fitting. The smaller the z-score, the closer the bulk of the posterior is to the true parameter [48]. In contrast, large z-values may be indicative of overfitting and, or poor prior specifications [48].

To examine the influence of the likelihood function in relation to prior information, we computed the posterior contraction, *k*:

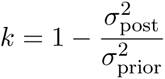

where 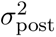 and 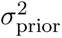 correspond to the variance of posterior and prior distributions, respectively. The *k* values close to zero indicate that data contain little information (i.e., rendering priors strongly informative). Conversely, values close to 1 indicate that data are much more informative than the prior [48].

We found that most of our model parameters and hyperparameters — were estimated with accuracy, precision, and identifiability, with the absolute posterior z-scores below three (Fig. S2). The posterior distributions for covariate coefficients, *β*, contracted by 86% on average, and at least 75%, compared to the prior distribution, meaning that the covariate coefficients were well-identified from data (Fig. S2). Although we used generic priors recommended by Stan [42], the *L_s_* parameters that define correlations among between-woman variation showed limited posterior contraction (i.e., ≤∼ 0.25), indicating that these parameters are poorly informed by data. As such, we refrain from making biological inferences about these correlations.

### S3: Predicted difference in community state type (CST) prevalence at various counterfactual scenarios

Our counterfactual simulations predicted that alcohol consumption and the number of partners are factors that impact the population-level outcome in terms of the prevalence of different community state types. The full list of comparisons is available in Fig. S3.

**Figure S2:**
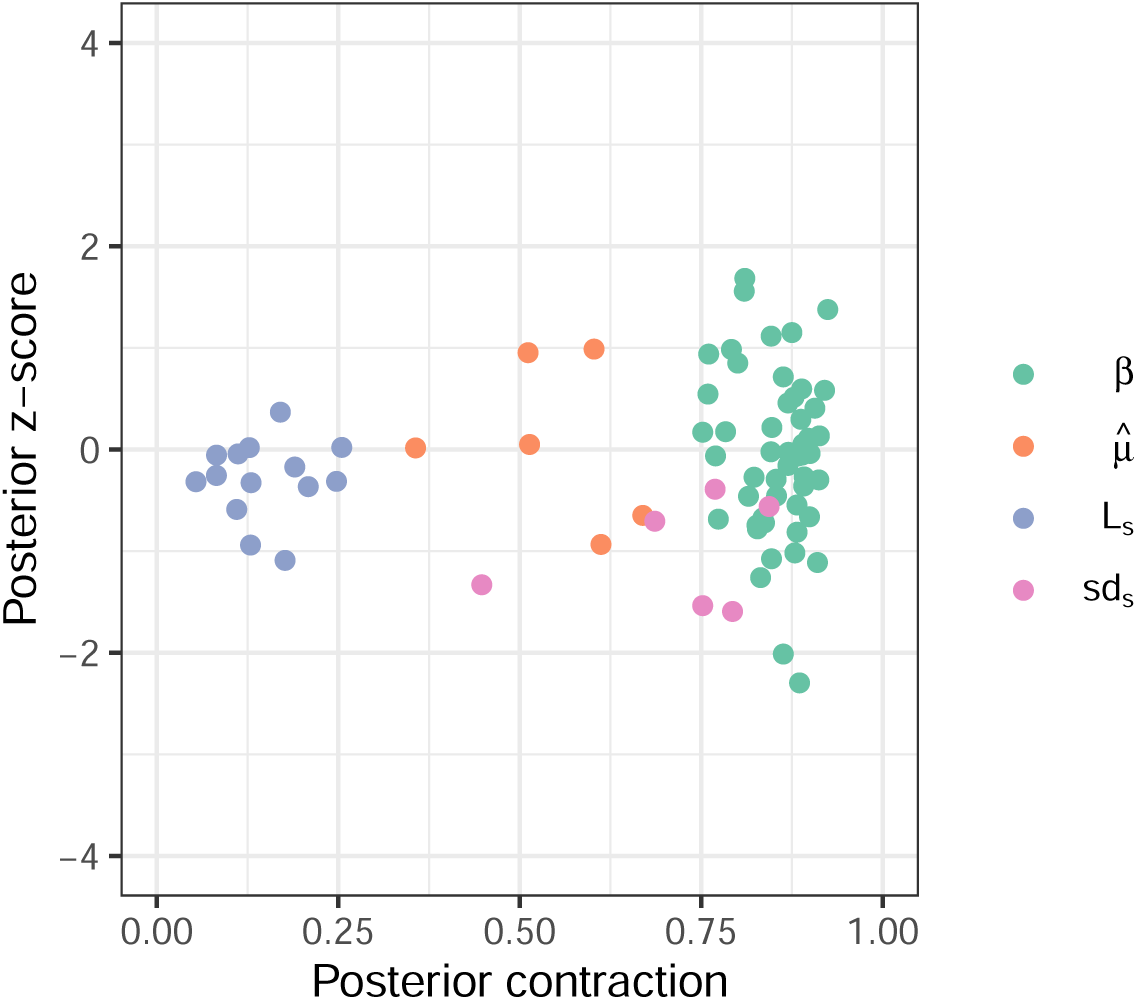
Accuracy, precision and identifiability of estimated parameters. Posterior z-score (y-axis) measures how closely the posterior recovers the parameters of the true data-generating process and posterior contraction (x-axis) evaluates the influence of the likelihood function over the prior, respectively. Smaller absolute posterior z-scores indicate that the posterior accurately recovers the parameters of the data-generating process: the absolute value beyond three to four may indicate substantial bias [48]. The posterior contraction values close to one indicate that data are much more informative than the prior. The estimated parameters are represented by a filled dot.

**Figure S3:**
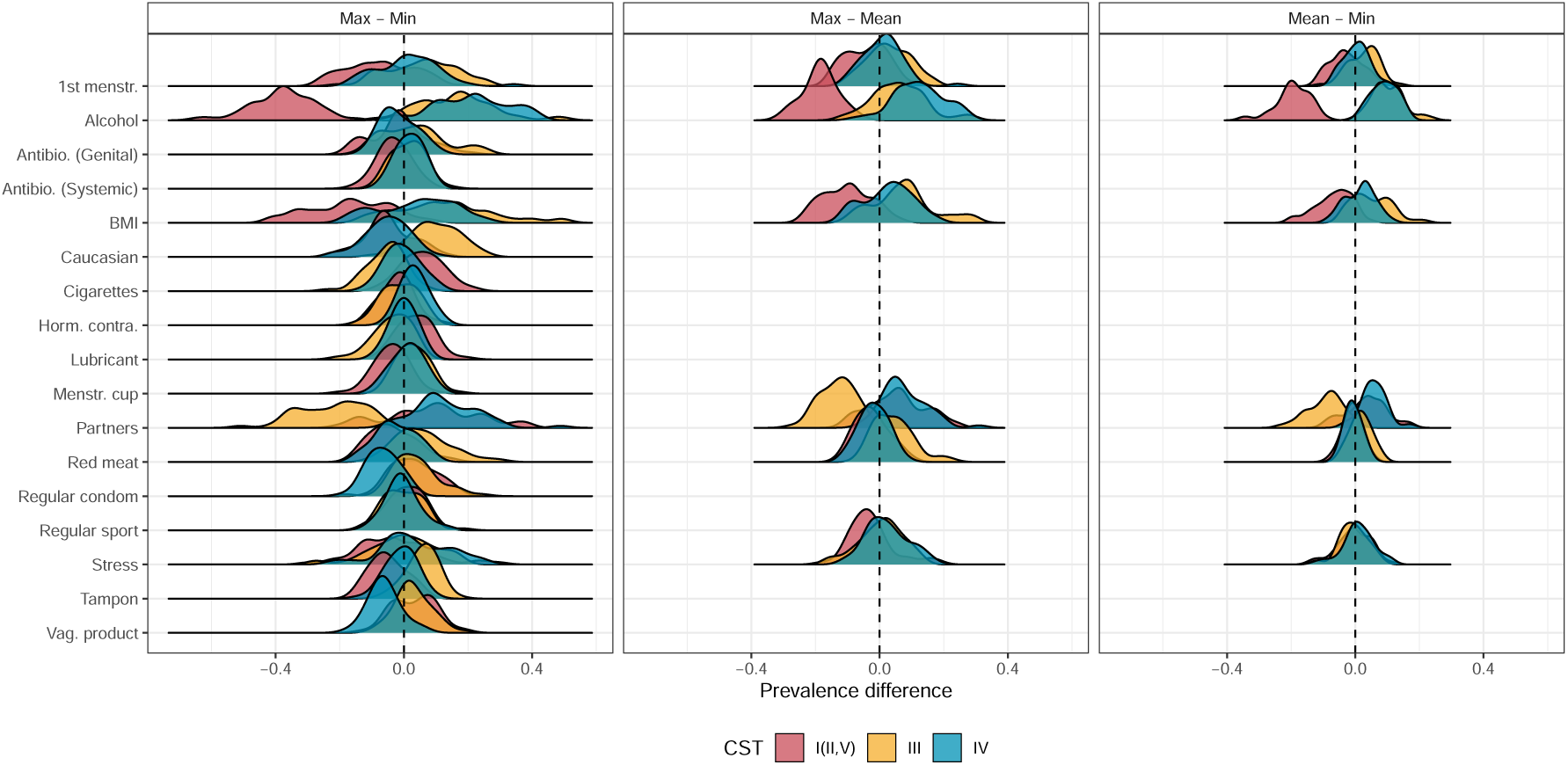
Difference in community state type (CST) prevalence at predicted various counterfactual scenarios. The differences were calculated from posterior samples simulated at 0 and 1 for binary variables and at the population maximum and minimum values recorded by the PAPCLEAR for continuous variables (left panel). Additional differences were computed between the population maximum and mean (middle panel) and the population mean and minimum for continuous variables (right panel). Parameter descriptions are found in Materials and Methods.

